# The gender gap in adolescent mental health: a cross-national investigation of 566,827 adolescents across 73 countries

**DOI:** 10.1101/2020.06.12.20129312

**Authors:** Olympia Campbell, David Bann, Praveetha Patalay

**Affiliations:** University College London, UK

## Abstract

Mental ill-health is a leading cause of disease burden worldwide. While women suffer from greater levels of mental health disorders, it remains unclear whether this gender gap differs systematically across regions and/or countries, or across the different dimensions of mental health. We analysed 2018 data from 566,827 adolescents across 73 countries for 4 mental health outcomes: psychological distress, life satisfaction, eudaemonia, and hedonia. We examine average gender differences and distributions for each of these outcomes as well as country-level associations between each outcome and purported determinants at the country level: wealth (GDP per capita), inequality (Gini index), and societal indicators of gender inequality (GII, GGGI, and GSNI). We report four main results: 1) The gender gap in mental health in adolescence is largely ubiquitous cross-culturally, with girls having worse average mental health; 2) There is considerable cross-national heterogeneity in the size of the gender gap, with the direction reversed in a minority of countries; 3) Higher GDP per capita is associated with worse average mental health and a larger gender gap across all mental health outcomes; and 4) more gender equal countries have larger gender gaps across all mental health outcomes. Taken together, our findings suggest that while the gender gap appears largely ubiquitous, its size differs considerably by region, country, and dimension of mental health. Findings point to the hitherto unrealised complex nature of gender disparities in mental health and possible incongruence between expectations and reality in high gender equal countries.

## Introduction

Mental ill-health is a leading cause of disease burden globally (1, 2), and in most individuals is first experienced in childhood (3), leading to a growing policy interest in improving adolescent mental health (4). During childhood and adolescence girls tend to report substantially worse internalising mental health than boys and this gender gap increases with age during adolescence (5–11). This may contribute to the disproportionately higher prevalence of common mental health disorders in adult women worldwide (12). It is important to document and understand cross-national differences in mental health with a focus on the gender gap: doing so may help identify countries with successful cultures and/or policies which could be implemented more broadly to reduce the gender mental health gap.

Despite evidence documenting a gender difference in adolescent mental health, it remains poorly understood. First, existing evidence is largely from a small number of high-income Western countries (7, 10, 13–17) and caution must be taken when generalizing their findings to non-Western, middle and low-income countries (18). Second, studies typically use only one measure of mental health; yet it is a multidimensional concept (19). As defined by the WHO (20), mental health is not simply the absence of mental illness but also a state of wellbeing and lies along a continuum from ill-health to positive mental health or wellbeing. It is constituted of several weakly correlated dimensions (21) including psychological distress, life satisfaction, hedonia (positive affect) and eudaemonia (the experience of purpose and meaning in life) (19). Third, most studies examine average differences in mental health between countries and genders, and do not explicitly examine its distribution. Understanding in which part of the population distribution average differences emerge may be useful to aid understanding of the nature of the gender gap and potential policy targets (22) – for instance, average gender differences may be due to a particularly high frequency of females at the severe end of the spectrum or due to differences across the entire distribution.

Cross-national comparisons can also identify factors at the country-level which are associated with mental health. Poverty is considered an established risk factor for worse mental health (23–25) and more unequal countries tend to have worse average mental health outcomes(16, 26–28). It is unknown however how wealth or income inequality are associated with the gender gap in mental health, and whether this differs by dimension of mental health — life satisfaction questions for example typically correlate more strongly with economic factors than affect-related questions (29).

Existing research on the association between gender equality and mental health largely yields inconsistent findings with studies demonstrating no association (14), stronger positive associations with both male mental health (25) and female mental health (30), and both smaller (7, 30) and larger mental health gender gaps (25, 31). Tesch-Romer et. al. (32) find that the association between gender equality and the adult mental health gender gap varies with the cultural attitudes of gender equality. Where over 50% agree with the statement ‘men have more of a right to work than a woman’, the mental health gender gap is larger with greater gender equality, but where less than 50% agree, the gap is smaller in countries with greater gender equality. Few studies, to our knowledge, have 1) explicitly examined the relationship between gender equality and the mental health gap in adolescents, 2) investigated the adolescent gender gap in a broad sample of countries including low- and middle-income countries and, 3) focused on multiple indicators of mental health.

Using a large cross-national dataset from 73 countries and economies and spanning a range of income groups, we aimed to 1) describe the gender gap across different measures of mental health (life satisfaction, psychological distress, hedonia, eudaemonia) in terms of both average and distributional differences, and 2) investigate the correlations of macro-level economic and gender equality indicators with wellbeing in boys and girls to better understand the gender mental health gap in adolescents. We hypothesised that: A) girls will have worse average mental health than boys across all outcomes; and B) that increased GDP and lower income inequality will be associated with better mental health outcomes for both genders; C) Higher gender equality will be associated with better mental health outcomes for both genders and a smaller mental health gap.

## Methods

### Participants

We used data from the 2018 Programme for International Student Assessment (PISA) (33). PISA is a multi-country cross-sectional study that surveys students at age 15 on their educational attainment and characteristics of their life (34). In total 73 countries and participating economies were included, containing 566,827 students (49.8% girls and 50.2% boys), representing around 28 million students. Countries excluded were: Singapore; Norway; New Zealand; and Israel as they did not collect the mental health measures. Subsamples that were not nationally representative were dropped, such as China. In order to investigate regional patterns, countries were grouped by region according to the World Health Organisation’s groupings (Table S1, see for example: https://www.who.int/choice/demography/euroregion/en/). The countries sampled cover a number of regions: North and South America; Europe; Eastern Mediterranean; South East Asia; and the Western Pacific Region.

### Measures

#### Outcome variables

Life satisfaction, psychological distress, hedonia and eudaemonia (35) were all measured in PISA 2018. Life satisfaction was measured by the question: “on a scale of 0-10, overall, how satisfied are you with your life as a whole these days?”, with 0 meaning not at all satisfied and 10 meaning completely satisfied. Psychological distress was assessed with responses to how often adolescents felt sad, miserable, scared, and afraid on a scale of never, rarely, sometimes, and always. Answers were scored 1-4 and summed to give an overall score ranging from 4-16. Hedonia was assessed with responses (never to always) to how often adolescents felt happy, lively, proud, joyful, and cheerful. Answers were summed to give an overall score ranging from 5-20. Eudaemonic wellbeing was measured by asking students how much they agreed on a scale of strongly disagree, disagree, agree, and strongly agree to the following statements: “my life has clear meaning or purpose”; “I have discovered a satisfactory meaning in life”; and “I have a clear sense of what gives meaning to my life”. The answers were scored and summed to give an overall score ranging from 3-12. In order to be able to compare scales each outcome was z-score standardised to have a mean of 0 and a variance of 1. Findings did not differ when examined in the original scales (data available upon request). Invariance testing showed that measures where invariant by gender, region and gender x region (Table S2). Original items can be found in the student questionnaire (33).

All questions were translated into the languages of participating countries by two independent linguists and then reconciled by a third to ensure consistent meaning in all countries. Further information can be found in the PISA technical report (34).

#### Gender

Gender was measured by students responding to the question “are you female or male?” coded 1 for girl and 0 for boy.

#### National Level Characteristics

Measures of gross domestic product (GDP) per capita and income inequality (Gini) were taken from the World Bank dataset. GDP per capita is the total economic output of a country divided by its population and is an estimate of prosperity. The Gini index is a measure of how unequal the income distribution is and ranges from 0, representing perfect equality, to 100 representing perfect inequality.

Three measures of gender equality were used in this study: the Gender Inequality Index (GII) and the newly created Gender Social Norms Index (GSNI) derived from the World Values Survey, both produced by the UNDP; and the Global Gender Gap Index (GGGI), produced by the World Economic Forum. Whilst all three use the same themes of education, health, political and economic participation they use different indicators to make these up (Table S3 for a summary of indicators). The main difference between the GII and the GGGI is that the GII is calculated in order to measure the loss in human development from gender inequality (see http://hdr.undp.org/sites/default/files/hdr2019_technical_notes.pdf). In contrast, the GGGI aims to separate gender equality from the country’s level of development by rewarding or penalizing countries based on the size of the gender gap in a particular resource regardless of the overall level of said resource (36). The GSNI is different from the other two as it tries to capture social norms through the proportion of people that agree or disagree with a particular statement, for example, “men make better political leaders than women do”. This allows us to test whether cultural attitudes towards gender equality are particularly important in terms of mental health outcomes.

### Analysis

We calculate country-level average differences for each standardised measure of mental health by calculating the weighted male and female mean for each country and then subtracting female average from male. Weighted means were calculated using the R package intsvy (37) designed to use the PISA provided weights and to take into account the two-stage sample design. Meta-analyses using the I^2^ statistic were performed to test heterogeneity in the gender differences between regions. The I^2^ statistic quantifies the percentage of total variation across nations due to heterogeneity rather than chance (38). To examine the distributions of mental health outcomes across the sample, weighted frequency histograms were plotted for each country for each outcome.

To explore the association of country-level factors on mental health outcomes, we estimated Pearson’s correlations (r) and plotted the relationships between the average score for each gender by country against the 5 country-level indicators: GDP per capita, Gini, GII, GSNI, GGGI. We use multi-level linear regression in order to estimate the between country variation in different mental health outcomes and to formally statistically investigate the associations between mental health, gender and country-level factors. Using weight scaling method A proposed by Asparouhov (39) and Carle (40) we adjust the final student weights by the number of individuals in each cluster divided by the sum of the sampling weights in each cluster (see (40), Appendix B), in order to estimate multi-level models. In these regressions, we use a single indicator of gender equality – GGGI – to avoid multicollinearity with other equality measures (Table S4).

## Results

### Do girls have worse average mental health than boys across all outcomes?

On average, girls have worse mental health across all indicators (Table 1). Life satisfaction and psychological distress have the largest mean differences between the sexes, 0.41 (0.33 s.d) and −1.1 (0.34 s.d) respectively, whereas hedonia and eudaemonia have smaller gender gaps, 0.10 (0.39 s.d) and 0.15 (0.27 s.d) respectively. The correlation matrix shows that individual-level correlations between mental health outcomes are weak-moderate - none reach 0.5 (Table 1, top half). The country-level correlations between the gender gaps (Table 1, bottom half) are all greater than 0.5 indicating that countries with large gender gaps in one outcome are likely to have large gender gaps in others.

**Table 1:** Descriptive statistics for all mental health outcomes showing the mean (and standard deviation and the individual-level and country-level correlations. Both unstandardised and standardised mean country gender gap are shown. Note that a positive gender gap indicates worse outcomes for girls apart from for psychological distress where a negative gender gap indicates worse outcomes for girls. *the non-shaded top half of the correlation matrices contains individual-level correlations between mental health outcomes. The shaded bottom half contains country-level correlations between the average gender gaps in mental health outcomes.

| Outcomes | Average Mental Health Scores |  |  |  | Correlations (r)* |  |  |  |
| --- | --- | --- | --- | --- | --- | --- | --- | --- |
|  | Males (SD) | Females (SD) | Unstandardised Gender Gap (SD) | Standardised Gender Gap (SD) | Life Satisfaction | Psychological Distress | Hedonia | Eudaemonia |
| **Life Satisfaction** | 7.3 (2.5) | 6.9 (2.5) | 0.41 (0.33) | 0.16 (0.13) |  | -0.34 | 0.49 | 0.40 |
| **Psychological Distress** | 9.1 (2.3) | 10.0 (2.1) | -1.1 (0.34) | -0.46 (0.14) | -0.67 |  | -0.23 | -0.21 |
| **Hedonia** | 16.2 (2.7) | 16.1 (2.6) | 0.10 (0.39) | 0.04 (0.14) | 0.69 | -0.53 |  | 0.41 |
| **Eudaemonia** | 8.8 (2.1) | 8.7 (2.0) | 0.15 (0.27) | 0.07 (0.12) | 0.79 | -0.54 | 0.53 |  |

In most countries girls have worse life satisfaction, and in all countries girls report more psychological distress than boys (Fig. 1). Hedonia and eudaemonia show greater cross-cultural variation with some countries exhibiting worse average outcomes for boys, such as Jordan and Saudi Arabia (Fig. 1). Some regional patterns emerge; wealthier European nations consistently have worse average mental health for girls across all outcomes apart from hedonia; the Eastern Mediterranean countries consistently have some of the smallest gender gaps, and for hedonia and eudaemonia have better average outcomes for girls. Particular countries consistently have some of the largest gender gaps in mental health, including Sweden, Finland, Slovenia and South Korea. For each outcome there was strong evidence for heterogeneity in the gender differences - both within and between regions with I^2^ > 95% for all outcomes, p <0.001 (Fig S1).

**Figure 1.**
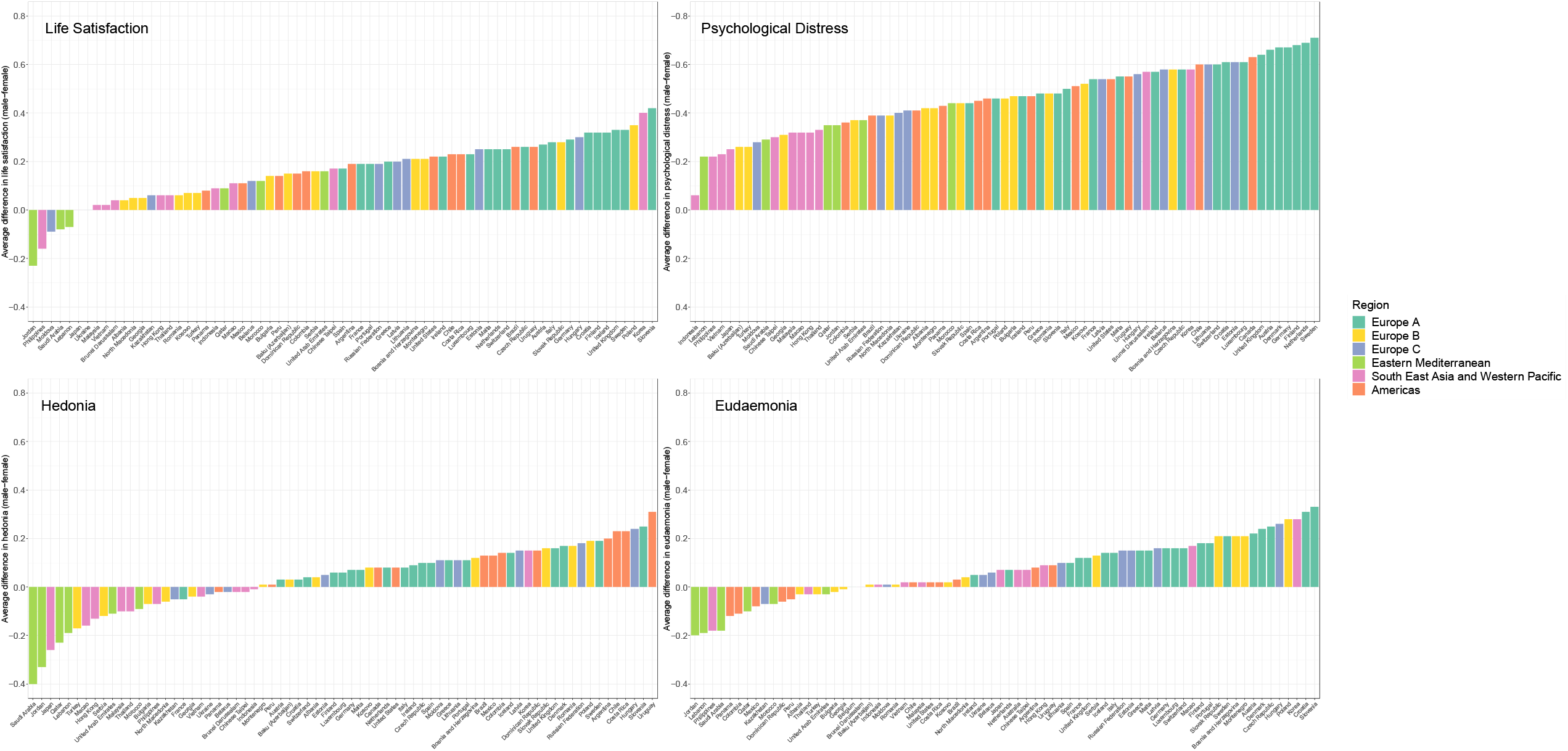
Average standardised gender difference (male – female) in mental health across each outcome by country and coloured by region. Average gender difference in mental health outcomes (life satisfaction, psychological distress, hedonia, and eudaemonia) for each country coloured by region. Gender difference is calculated by subtracting the female from the male mean. The y-axis of the psychological distress scale is reversed to allow visual comparison with the other mental health outcomes as a more negative difference for psychological distress indicates worse outcomes for girls.

### Distributions

Examination of distributions revealed that average gender differences in life satisfaction were driven by different parts of the wellbeing distribution; boys have higher upper values of life satisfaction (9/10 out of 10) (Fig. S2); while for psychological distress (Fig. 2) the female distribution is overall shifted to the right, indicating a higher frequency of feelings of distress in girls across the spectrum. Hedonia is also largely left skewed (Fig. S3) and the distributional gender differences are less pronounced. Eudaemonia peaks at 9 for both boys and girls in most of the countries and the gender difference looks uniform across the distribution (Fig. S4). Thus, despite different overall distributions, the mental health gender gap remains, although where the gap appears in the distribution differs by outcome.

**Figure 2:**
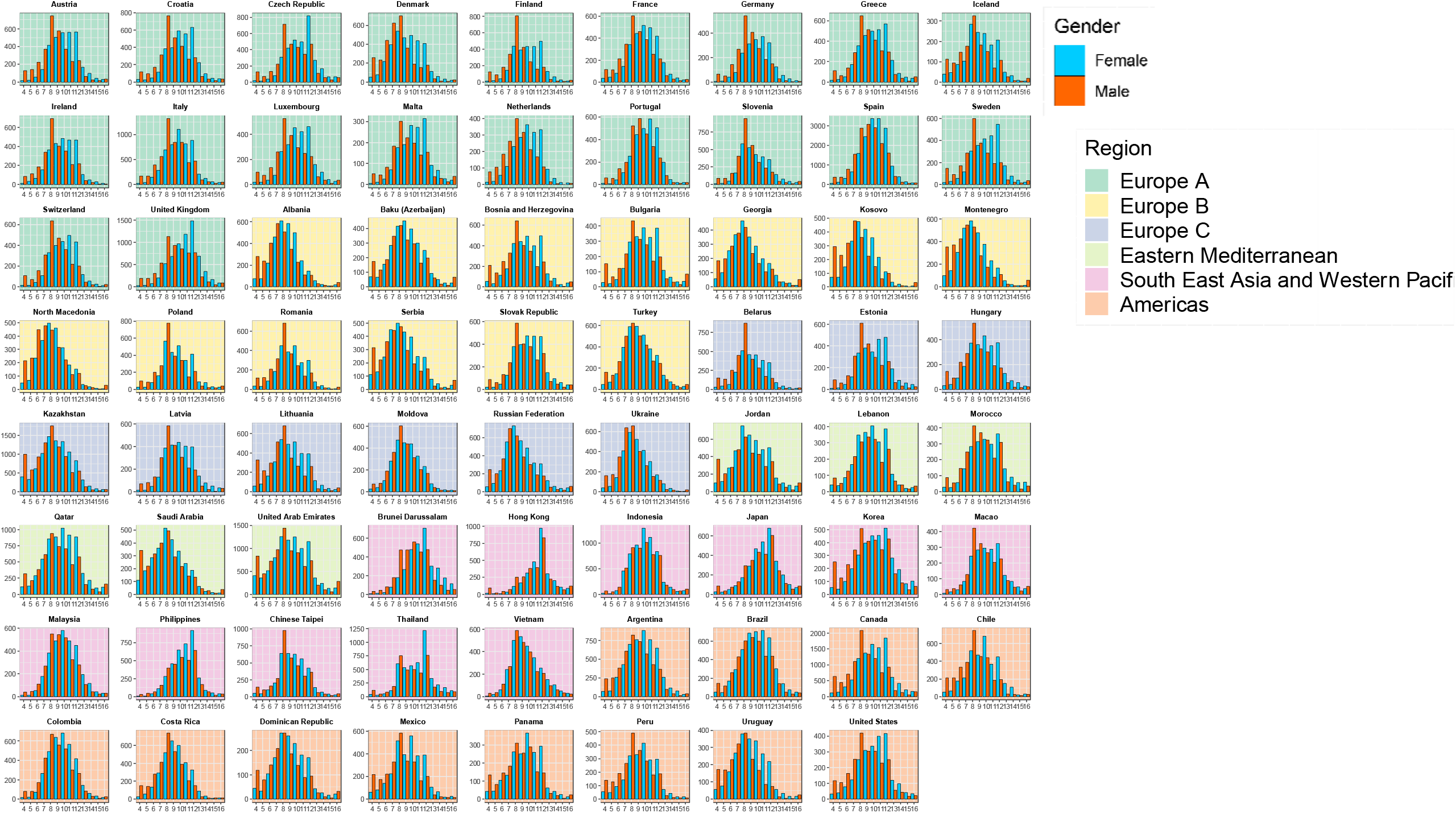
Distributions of psychological distress for males and females by country and region.

### Country Level Associations

The proportion of total variance attributable to differences between countries was estimated to be 5.6% for life satisfaction, hedonia and eudaemonia and 7.3% for psychological distress (using the variance partition coefficient from the baseline multi-level model (Table S5 Model A). Overall, the final model explains 35.7% of the between country variance in life satisfaction, 8.2% in psychological distress, 16.1% in hedonia, and 46.4% in eudaemonia. Figures 3 and S5 present the associations between the country-level indicators and each mental health outcome by gender.

**Figure 3:**
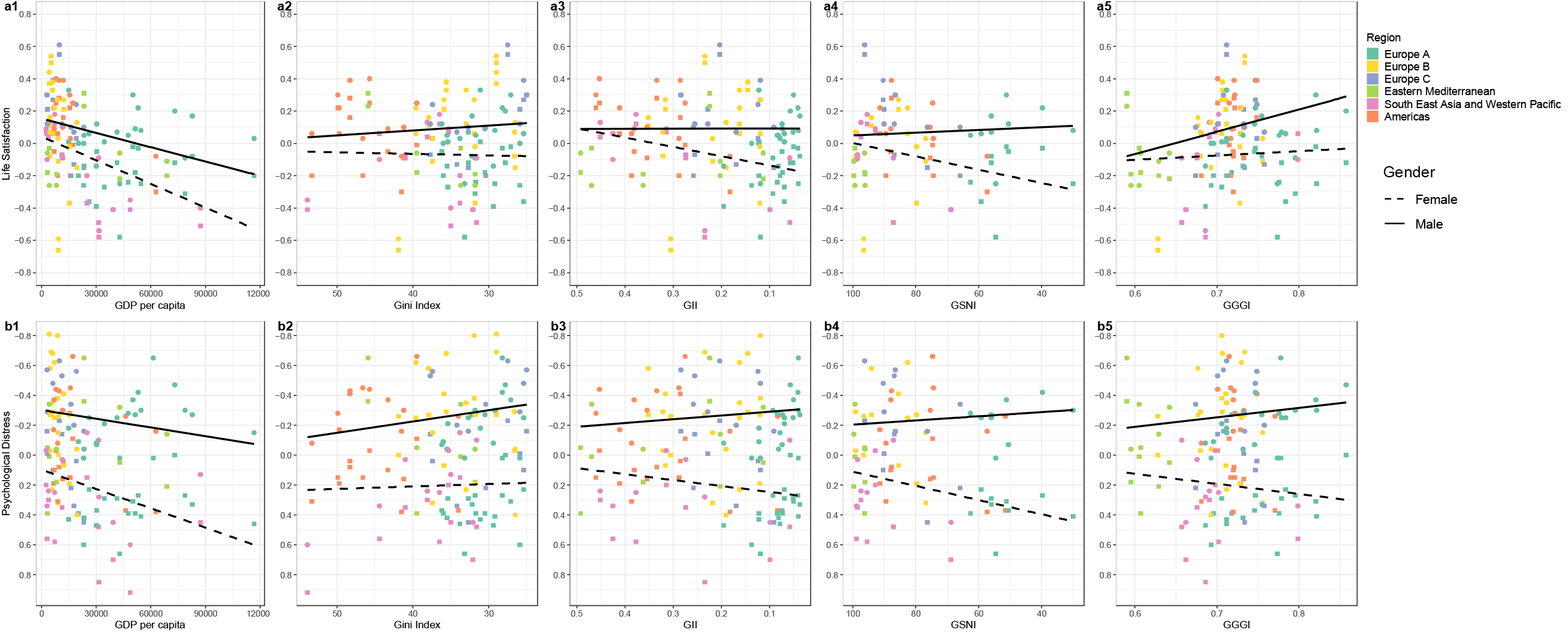
Associations of country-level economic and gender equality indicators with average life satisfaction and psychological distress by gender. Country-level associations of economic indicators (GDP per capita and Gini) and gender equality indicators (GII, GSNI, and GGGI) with average standardised life satisfaction (a1-5) and psychological distress (b1-5) for females and males and coloured by region. The GII, GSNI and Gini scales are reversed so that all x-axis run from less equal to more equal. The psychological distress scale is reversed so that a negative relationship indicates worse mental health across all outcomes. A larger distance between the regression lines indicates a larger gender gap. Abbreviations: Gini = income inequality, GII = gender inequality index, GSNI = gender social norms index, GGGI = global gender gap index.

### Is higher GDP and lower income inequality associated with better mental health outcomes for both genders?

Higher GDP per capita was associated with lower life satisfaction (β −0.035 [0.012sd]), hedonia (−0.027 [0.013sd]) and eudaemonia (−0.037 [0.01sd]) and higher psychological distress (0.033 [0.014sd]) for both boys and girls (Fig. 3, Table S5). For all outcomes (except hedonia) the gender gap was larger for wealthier nations mainly driven by steeper slopes for females (Fig. 3 and S5).

Higher income inequality was associated with slightly lower life satisfaction for boys and slightly higher life satisfaction for girls and thus a slightly smaller gender gap in more unequal countries (Fig. 3: a2). Higher income inequality was associated with marginally more psychological distress for both genders (0.0006 [0.005sd], but this association is slightly stronger for boys than girls and thus more equal countries have larger gender gaps (Fig. 3: b2). By contrast, lower income inequality was associated with lower hedonia and eudaemonia and slightly larger gender gaps (Fig.S5: a2 & b2). Thus, while more equal countries have larger gender gaps across all outcomes the direction of association between Gini and mental health differs by outcome.

### Is higher gender equality associated with better mental health for both genders and a smaller gender gap?

More gender equality was associated with a larger gender gap across all mental health outcomes (Fig. 3 and S5; Table S5). The processes underlying this larger gender gap differed by outcome. The larger gap in life satisfaction and psychological distress was mostly driven by positive correlations with male mental health but negative correlations with female mental health, apart from the association between GGGI and female life satisfaction which was weakly positive (Fig. 3: a5). The widening gap in hedonia and eudaemonia was mostly due to stronger negative correlations with female mental health and weaker negative correlations with male mental health, apart from the association between GGGI and male hedonia which was positive (0.20 [0.059sd] Table S5; Fig S5). The interaction terms between GGGI and gender are large so there is fairly strong evidence that the effect for gender differs with GGGI for all mental health outcomes, apart eudaemonia (Table S5).

## Discussion

Across four mental health outcomes - life satisfaction, psychological distress, hedonia, and eudaemonia - we find that girls typically had worse mental health than boys. Whilst there is considerable cross-cultural variation in the size of this average difference, it appears largely ubiquitous in this global sample - particularly for life satisfaction and psychological distress. Perhaps counterintuitively, richer European countries including the Scandinavian nations, such as Sweden and Finland, have some of the largest gender gaps in mental health. By contrast, countries with worse society gender equality scores – such as Jordan, Saudi Arabia, and Lebanon - have some of the smallest gender gaps and the direction of the gap is sometimes reversed (with boys having worse mental health). The outcomes vary in their distributions and where in the distribution the gender gap appears. Life satisfaction shows a marked gender difference at the end of the scale with boys more often scoring themselves 10 out of 10, while gender differences were found across the entire distribution for psychological distress.

Higher GDP per capita was associated with a larger gender gap, albeit the magnitude of effect was small. This contrasts with other findings where a positive relationship between GDP and adolescent wellbeing has been found (7), and this may be due to our inclusion of a wider range of countries beyond rich Western economies. The Easterlin paradox of increasing per capita wealth not associating with increasing wellbeing is well known (41) — once basic requirements are met, material desires often increase with increasing incomes so that one is never completely satisfied (29). This however does not completely explain the negative association with mental health we found in both genders, or the larger mental health gender gap in richer countries. In contrast to previous literature (26) we do not find a consistent relationship between income inequality and mental health outcomes, although it is associated with a wider gender gap in all cases. It could be the case that income inequality is not particularly important amongst adolescents, and that any effects if present, are more manifest in adulthood.

More gender equal countries had larger gender gaps across all outcomes examined. Whilst the nature of the associations between gender equality and mental health were inconsistent across outcomes it was striking that where the association was positive, it was particularly strong for males. This is in contrast to previous findings that show an equivalent positive relationship between gender equality and life satisfaction in boys and girls (17). Whilst previous work has shown that social norms of gender equality may be particularly important for mental health outcomes (32) it is unclear if the multiple available gender equality indicators we used fully capture this. The newly created gender social norms index (GSNI), despite attempting to capture the distinct attitudinal aspects of gender equality, does not appear to measure gender equality in a qualitatively different way than the GII. By contrast the GGGI captures a greater detail of gender equality by including more indicators, making it more granular, whilst also separating itself from a country’s level of development.

Our results present a complex picture for the relationship between gender equality and the adolescent gender mental health gap. The movement towards gender equality is a fairly recent development, with the UN Convention on the Elimination of all Forms of Discrimination Against Women (CEDAW) only being instituted in 1981. Graham and Pettinato (42) coined the term ‘frustrated achievers’ to describe individuals that experience improvements in wealth but report negative perceived past mobility and lower happiness, as a result of still facing discriminatory practices and barriers to their continued ascent. In terms of women, whilst gains have been made, there remain many barriers to full equality that may explain part of our association between gender equality and worse female mental health, or only very slightly better female mental health in the case of life satisfaction. Similarly, expectations of equality may rise faster than actual experience of equality and this may result in worse mental health as women are not able to realise their goals. Another characteristic of upwardly mobile groups is that their reference categories for social comparison are usually beyond their original cohort (41). Thus, women or girls attempting to achieve the same successes as men and boys will look to them as their reference group and this may highlight the inequalities between them, producing lower life satisfaction and mental health, while in less gender equal countries reference groups might be limited to their own sex (31).

In more gender equal countries girls and women are now faced with a double burden of balancing both increased economic and political participation as well as the traditional female responsibilities and norms. In countries with lower gender equality women’s roles are more fixed, whereas in more gender equal countries they are less prescribed, leading to potential conflict between roles, which may affect mental health (43). Adolescence and puberty marks a particular period of changing identity (44) including developing conceptions of what it means to be a man or a woman (45). This can be particularly stressful when the norms of femininity potentially contradict with the norms of gender equality and attempting to balance the two may be additionally difficult. Indeed, changing norms of female education and economic participation can increase educational stress and psychological distress for girls whilst they are still burdened with traditional anxieties related to maintaining a female identity and appearance (9).

### Limitations

Firstly, our study relies exclusively on cross-sectional cross-country correlations; thus, we cannot make any strong conclusions regarding the causal pathways involved. However, cross-country comparisons are necessary to elucidate risk factors that operate at the population level (46), such as indicators of gender and income inequality. Secondly, whilst we cannot exclude cultural differences on likert scale responses, such as positivity biases, that may confound cross-country differences (47) invariance testing of the measures indicated that the measures behaved similarly across gender and region. Thirdly, the gender gap itself may partly be a product of reporting bias – with boys being less willing to report negative mental health than girls. However, self-reports are necessary to measure mental health and wellbeing, and the extent and distributions of the gender gap being different across mental health outcomes suggests reporting biases might not be the only explanation. Fourthly, there could be systematic differences across genders in school attendance amongst the countries in our sample that could potentially bias comparison of gender gaps across countries. However, investigation of the gender ratio in secondary enrolment (obtained from the GGGI) suggests that there are not large differences in our sample. The female to male ratio in secondary enrolment ranges from 0.9 to 1.1 for our whole sample, apart from Germany (0.89), the Philippines (1.19) and Qatar (1.25). Lastly, our measure of gender was binary in nature and does not allow investigation of non-binary gender identities on mental health.

### Conclusion

Our findings demonstrate that overall girls have worse mental health than boys, but the direction and size of the gender gap and distribution varies across a range of mental health outcomes and a large sample of countries. Wealthier and more gender-equal countries, contrary to expectation, have larger mental health gender gaps. For life satisfaction and psychological distress, this was driven by negative associations in females but positive associations in males. Findings point to the hitherto unrealised complex nature of gender disparities in mental health and possible incongruence between expectations and reality in more gender equal countries.

## Data Availability

All data used is available on the PISA website and World Bank, UNDP and Wolrd Economic Forum websites.

https://www.oecd.org/pisa/data/2018database/

## Acknowledgments

We thank the OECD for collecting and providing the PISA data. Olympia Campbell thanks the Economic and Social Research Council and the European Research Council for funding her PhD.

