## Supplementary Materials for "The gender gap in adolescent mental health: a cross-national investigation of 566,827 adolescents across 73 countries"

**Table S1: Regional groupings**

| EUR A | EUR B | EUR C | Eastern Mediterranean (EMR) | South East Asia/ Western Pacific (SEA WPR) | Americas (AMR) |
|---|---|---|---|---|---|
| Austria | Albania | Belarus | Jordan | Australia | Argentina |
| Belgium | Baku (Azerbaijan) | Estonia | Lebanon | Brunei Darussalam | Brazil |
| Croatia | Bosnia and Herzegovina | Hungary | Morocco | Chinese Taipei | Canada |
| Czech Republic | Bulgaria | Kazakhstan | Qatar | Hong Kong | Chile |
| Denmark | Georgia | Latvia | Saudi Arabia | Indonesia | Colombia |
| Finland | Kosovo | Lithuania | United Arab Emirates | Japan | Costa Rica |
| France | Montenegro | Moldova | | Korea (Republic of) | Dominican Republic |
| Germany | Poland | Russian Federation | | Macao | Mexico |
| Greece | Romania | Ukraine | | Malaysia | Panama |
| Iceland | Serbia | | | Philippines | Peru |
| Ireland | Slovak Republic | | | Vietnam | United States |
| Italy | Turkey | | | Thailand | Uruguay |
| Luxembourg | North Macedonia | | | | |
| Malta | | | | | |
| Netherlands | | | | | |
| Portugal | | | | | |
| Slovenia | | | | | |
| Spain | | | | | |
| Sweden | | | | | |
| Switzerland | | | | | |
| United Kingdom | | | | | |

Regional groupings based on World Health Organisation categories. South East Asia and Western Pacific groups were combined, and Kosovo, Serbia and Montenegro were all classified as Europe B as the WHO does not include them in their classifications

**Table S2: Measurement Invariance for Psychological Distress, Hedonia, and Eudaemonia**

| | Configural | Metric | Scalar |
|---|---|---|---|
| **Gender (2 groups)** | | | |
| RMSEA[1] | 0.077 | 0.074 | 0.071 |
| SRMR[2] | 0.042 | 0.043 | 0.043 |
| CFI[3] | 0.969 | 0.969 | 0.967 |
| TLI[4] | 0.960 | 0.963 | 0.967 |
| **Region (6 groups)** | | | |
| RMSEA[1] | 0.081 | 0.078 | 0.079 |
| SRMR[2] | 0.045 | 0.045 | 0.048 |
| CFI[3] | 0.967 | 0.966 | 0.954 |
| TLI[4] | 0.958 | 0.961 | 0.960 |
| **Region X Gender (12 groups)** | | | |
| RMSEA[1] | 0.080 | 0.077 | 0.081 |
| SRMR[2] | 0.044 | 0.046 | 0.049 |
| CFI[3] | 0.968 | 0.965 | 0.950 |
| TLI[4] | 0.959 | 0.961 | 0.958 |

**[1] Root Mean Square Error of Approximation**
**[2] Standardised Root Mean Square Residual**
**[3] Comparative Fit Index**
**[4] Tucker-Lewis Index**

We tested measurement invariance for the 3 outcome measures that had more than one item (the life satisfaction is a single item measure), by fitting a model where the items of each of the three scales loaded onto the respective scale. We investigated measurement invariance at three levels: configural, metric and scalar[1]. For the RMSEA and SRMR, a score closest to zero indicates good model fit, whereas for CFI it is closest to one. Rules of thumb for model comparison are as follows: for RMSEA invariance is met if a change is smaller than 0.015, for SRMR if it is smaller than 0.03 and for CFI if it is smaller than 0.01[2]. We investigated three measurement invariance by three factors: gender (2 groups: male and female), region (6 groups: Europe A, Europe B, Europe C, Eastern Mediterranean, South East Asia/Western Pacific, and Americas) and genderxregion (12 groups: each region by sex). As can be seen from Table S2 above, the measurement invariance conditions were adequately met.  We are therefore satisfied that the group invariance assumption is met.

[1] Putnick & Bornstein (2016) Measurement invariance conventions and reporting: The state of the art and future directions for psychological research. Developmental review. 41:71-90. doi: 10.1016/j.dr.2016.06.004

[2] Chen, F. F. (2007). Sensitivity of goodness of fit indexes to lack of measurement invariance. *Structural Equation Modeling*, *14*(3), 464–504. https://doi.org/10.1080/10705510701301834

**Table S3: Comparison of the different gender equality measures and the indicators that make up each dimension.**

| *Index* | *Dimension* | | | |
|---|---|---|---|---|
| | **Political** | **Educational** | **Economic** | **Health** |
| **Gender Inequality Index** | 1. Female and male shares of parliamentary seats | 1. Female and male population with at least secondary education | 1. Female and male labour force participation rates | 1. Maternal mortality ratio<br>2. Adolescent birth rate |
| **Global Gender Gap Index** | 1. Ratio of female to male seats in parliament<br>2. Ratio of females to males at ministerial level<br>3. Ratio of number of years with a female head of state (last 50 years) to years with male head of state | 1. Ratio of female literacy rate to male<br>2. Ratio of female net primary enrolment rate to male<br>3. Ratio of female net secondary enrolment rate to male<br>4. Ratio of female gross tertiary enrolment ratio to male | 1. Ratio of female labour force participation to male<br>2. Wage equality between women and men for similar work<br>3. Ratio of female estimated earned income to male<br>4. Ratio of female legislators, senior officials and managers over male value<br>5. Ratio of female professional and technical workers to males | 1. Sex ratio at birth<br>2. Ratio of female healthy life expectancy to males |
| **Gender Social Norms Index** | 1. “men make better political leaders than women do”<br>2. “women have the same rights as men” | 1. “University is more important for a man than for a woman” | 1. “men should have more right to a job than women”<br>2. “men make better business executives than women do” | 1. Proxy for intimate partner violence<br>2. Proxy for reproductive rights |

The GGGI measures gaps in gender equality rather than levels. For example, countries will score higher or lower on the GGGI depending on the difference in higher education enrolment between men and women but not for the overall level of higher education in the country. This separates the GGGI from a country’s level of development to a greater extent than the GII.   The items in the GSNI are derived from the World Values Survey

**Table S4: Correlation matrix showing the country-level correlations between the country-level variables.**

| | GII | GGGI | GSNI | GDP per cap | Gini |
|---|---|---|---|---|---|
| **GII** | 1.000 | -0.56 | 0.82 | -0.79 | 0.60 |
| **GGGI** | | 1.000 | -0.65 | 0.48 | -0.20 |
| **GSNI** | | | 1.000 | -0.85 | 0.47 |
| **GDP per cap** | | | | 1.000 | -0.41 |
| **Gini** | | | | | 1.000 |

Abbreviations: GII = Gender Inequality Index; GGGI = Global Gender Gap Index; GSNI = Gender Social Norms Index; GDP per cap = Gross Domestic Product per capita. The GII and GSNI are more highly correlated with the economic indicators GDP per capita and Gini than the GGGI is.

**Table S5:** Multi-level models for each mental health outcome showing regression coefficients and between-country variance (VPC)

| | Life Satisfaction Coef (SE) | Psychological Distress Coef (SE) | Hedonia Coef (SE) | Eudaemonia Coef (SE) |
|---|---|---|---|---|
| **Model A: Baseline model** | | | | |
| ***Country VPC*** | 5.6% | 7.3% | 5.6% | 5.6% |
| **Model B: including gender** | | | | |
| **Female** | -0.16 *** | 0.46*** | -0.035* | -0.069*** |
| | (0.015) | (0.02) | (0.016) | (0.014) |
| ***Country VPC*** | 5.4% | 8.1% | 6.05% | 4.3% |
| **Model C: including country indicators** | | | | |
| **Female** | -0.17*** | 0.47*** | -0.048** | -0.074*** |
| | (0.017) | (0.017) | (0.017) | (0.016) |
| **GGGI * 10** | 0.17** | -0.019 | 0.17** | 0.048 |
| | (0.052) | (0.064) | (0.058) | (0.046) |
| **GDP per cap x $10^{-4}$** | -0.035 ** | 0.033* | -0.027* | -0.037*** |
| | (0.012) | (0.014) | (0.013) | (0.01) |
| **Gini** | -0.00021 | 0.0006 | 0.0095* | 0.0037 |
| | (0.0036) | (0.005) | (0.0041) | (0.0032) |
| ***Country VPC*** | 3.6% | 7.1% | 4.7% | 3.0% |
| **Model D : cross level interactions** | | | | |
| **Female** | 0.37 | -0.08 | 1.008 | 0.039 |
| | (0.23) | (0.22) | (0.24) | (0.21) |
| **GGGI x 10** | 0.18*** | -0.077 | 0.20** | 0.034 |
| | (0.051) | (0.068) | (0.059) | (0.046) |
| **GDP per capita x $10^{-4}$** | -0.033** | 0.015 | -0.027* | -0.04*** |
| | (0.011) | (0.015) | (0.013) | (0.01) |
| **Gini** | -0.0003 | 0.002 | 0.0097* | 0.0058 |
| | (0.003) | (0.005) | (0.0041) | (0.0032) |
| **GGGI x10 X Female** | -0.076* | 0.079 ** | -0.14*** | -0.05 |
| | (0.03) | (0.03) | (0.031) | (0.027) |
| **GDP per cap x $10^{-4}$ X Female** | -0.017* | 0.025*** | 0.0027 | -0.0089 |
| | (0.0068) | (0.006) | (0.007) | (0.006) |
| **Gini X Female** | 0.0014 | -0.002 | -0.00104 | 0.0076*** |
| | (0.0021) | (0.002) | (0.0022) | (0.0019) |
| ***Country VPC*** | 3.6% | 6.7% | 4.7% | 3.0% |

Regression coefficients with standard errors (SE) from multilevel models. Model A presents the baseline model in order to calculate the country variance partition coefficient (VPC). Model B includes only sex, Model C contains all country-level factors and Model D contains all cross-level interactions between sex female and country-level factors. Since the GGGI scale runs from 0-1 we multiply it by 10 so that the coefficient for GGGI represents a 0.1-point increase in the scale. GDP per capita is rescaled by dividing by 10,000, so that the coefficient represents the association with an increase of $1 \times 10^4$ GDP per capita. Note that higher values of Gini indicate greater income inequality and that a positive coefficient for psychological distress indicates worse mental health in contrast to the other outcomes. Only the GGGI as a measure of gender equality is used due to the high correlations between the GII and GSNI and the economic variables (Table S3). *$p<0.05$ **$p<0.01$ ***$p<0.001$

**Table S5: Regression coefficients with standard errors (SE) and variance components of multi-level models for life satisfaction**

| | **Model A: Baseline Model** Coef (SE) | **Model B: + Sex** Coef (SE) | **Model C: + country-level indicators** Coef (SE) | **Model D: + cross-level interactions** Coef (SE) |
|---|---|---|---|---|
| Intercept | -0.00046 (0.029) | 0.079 (0.028) ** | -1.06 (0.40)* | -1.1 (0.39)** |
| Sex female | | -0.16 (0.015) *** | -0.17 (0.017)*** | 0.37 (0.23) |
| GGGI x 10 | | | 0.17 (0.052)** | 0.18 (0.051)*** |
| GDP per capita x $10^{-4}$ | | | -0.035 (0.012) ** | -0.033 (0.011)** |
| Gini | | | -0.00021 (0.0036) | -0.0003 (0.003) |
| GGGI x 10 X sex female | | | | -0.076 (0.03)* |
| GDP per cap x $10^{-4}$ X sex female | | | | -0.017 (0.0068)* |
| Gini X sex female | | | | 0.0014 (0.0021) |
| **Variance Components** | | | | |
| Individual | 0.95 (0.97) | 0.94 (0.97) | 0.93 (0.96) | 0.93 (0.96) |
| National**:** Country | 0.056 (0.24) | 0.055 (0.24) | 0.035 (0.19) | 0.035 (0.19) |
| Sex Female | | 0.016 (0.12) | 0.017 (0.13) | 0.012 (0.11) |
| **VPC** | 5.6% | 5.4% | 3.6% | 3.6% |

Model A baseline model to illustrate the between country variance. Model B includes sex female. Model C includes all country-level variables. Model D includes all cross-level interactions with sex female. Since the GGGI scale runs from 0-1 we multiply it by 10 so that the coefficient for GGGI represents a 0.1-point increase in the scale. GDP per capita is rescaled by dividing by 10,000, so that the coefficient represents the association with an increase of 1 x $10^4$ GDP per capita. Variance partition coefficient (VPC) indicates the proportion of unexplained variance attributable to differences between countries. *p<0.05 **p<0.01 ***p<0.001

**Table S6: Regression coefficients with standard errors (SE) and variance components of multi-level models for psychological distress**

| | **Model A: Baseline Model** Coef (SE) | **Model B: + Sex** Coef (SE) | **Model C: + country-level indicators** Coef (SE) | **Model D: + cross-level interactions** Coef (SE) |
|---|---|---|---|---|
| Intercept | -0.019 (0.032) | -0.25 (0.03)*** | -0.24 (0.50) | 0.17 (0.53) |
| Sex female | | 0.46 (0.02) *** | 0.47 (0.017)*** | -0.08 (0.22) |
| GGGI x 10 | | | -0.019 (0.064) | -0.077 (0.068) |
| GDP per capita x $10^{-4}$ | | | 0.033 (0.014) * | 0.015 (0.015) |
| Gini | | | 0.0006 (0.005) | 0.002 (0.005) |
| GGGI x 10 X sex female | | | | 0.079 (0.03) ** |
| GDP per cap x $10^{-4}$ X sex female | | | | 0.025 (0.006) *** |
| Gini X sex female | | | | -0.002 (0.002) |
| **Variance Components** | | | | |
| Individual | 0.93 (0.96) | 0.87 (0.94) | 0.85 (0.92) | 0.87 (0.93) |
| National**:** Country | 0.073 (0.27) | 0.079 (0.28) | 0.067 (0.26) | 0.048 (0.22) |
| Sex Female | | 0.018 (0.14) | 0.019 (0.14) | 0.012 (0.11) |
| **VPC** | 7.3% | 8.1% | 7.1% | 6.7% |

Model A baseline model to illustrate the between country variance. Model B includes sex female. Model C includes all country-level variables. Model D includes all cross-level interactions with sex female. Since the GGGI scale runs from 0-1 we multiply it by 10 so that the coefficient for GGGI represents a 0.1-point increase in the scale. GDP per capita is rescaled by dividing by 10,000, so that the coefficient represents the association with an increase of 1 x $10^4$ GDP per capita. Variance partition coefficient (VPC) indicates the proportion of unexplained variance attributable to differences between countries. *p<0.05 **p<0.01 ***p<0.001

**Table S7: Regression coefficients with standard errors (SE) and variance components of multi-level models for Hedonia**

| | **Model A: Baseline Model** Coef (SE) | **Model B: + Sex** Coef (SE) | **Model C: + country-level indicators** Coef (SE) | **Model D: + cross-level interactions** Coef (SE) |
|---|---|---|---|---|
| Intercept | -0.012 (0.028) | 0.006 (0.03) | -1.5 (0.45) ** | -1.7 (0.45)*** |
| Sex female | | -0.035 (0.016) * | -0.048 (0.017)** | 1.008 (0.24)*** |
| GGGI x 10 | | | 0.17 (0.058) ** | 0.20 (0.059)** |
| GDP per capita x $10^{-4}$ | | | -0.027 (0.013) * | -0.027 (0.013)* |
| Gini | | | 0.0095 (0.0041)* | 0.0097 (0.0041)* |
| GGGI x 10 X sex female | | | | -0.14 (0.031)*** |
| GDP per cap x $10^{-4}$ X sex female | | | | 0.0027 (0.007) |
| Gini X sex female | | | | -0.00104 (0.0022) |
| **Variance Components** | | | | |
| Individual | 0.94 (0.97) | 0.94 (0.97) | 0.92 (0.96) | 0.92 (0.96) |
| National**:** Country | 0.056 (0.24) | 0.062 (0.25) | 0.045 (0.22) | 0.046 (0.22) |
| Sex Female | | 0.018 (0.14) | 0.018 (0.13) | 0.013 (0.11) |
| **VPC** | 5.6% | 6.05% | 4.7% | 4.7% |

Model A baseline model to illustrate the between country variance. Model B includes sex female. Model C includes all country-level variables. Model D includes all cross-level interactions with sex female. Since the GGGI scale runs from 0-1 we multiply it by 10 so that the coefficient for GGGI represents a 0.1-point increase in the scale. GDP per capita is rescaled by dividing by 10,000, so that the coefficient represents the association with an increase of 1 x $10^4$ GDP per capita. Variance partition coefficient (VPC) indicates the proportion of unexplained variance attributable to differences between countries. *p<0.05 **p<0.01 ***p<0.001

**Table S8: Regression coefficients with standard errors (SE) and variance components of multi-level models for Eudaemonia**

| | **Model A:** **Baseline Model** Coef (SE) | **Model B:** **+ Sex** Coef (SE) | **Model C:** **+ country-level indicators** Coef (SE) | **Model D:** **+ cross-level interactions** Coef (SE) |
|---|---|---|---|---|
| Intercept | -0.0024 (0.028) | 0.032 (0.024) | -0.34 (0.36) | -0.31 (0.36) |
| Sex female | | -0.069 (0.014) *** | -0.074 (0.016)*** | 0.039 (0.21) |
| GGGI x 10 | | | 0.048 (0.046) | 0.035 (0.047) |
| GDP per capita x $10^{-4}$ | | | -0.037 (0.01) *** | -0.04 (0.01)*** |
| Gini | | | 0.0037 (0.0032) | 0.0058 (0.0033) |
| GGGI x 10 X sex female | | | | -0.05 (0.027) |
| GDP per cap x $10^{-4}$ X sex female | | | | -0.0090 (0.006) |
| Gini X sex female | | | | 0.0076 (0.0019)*** |
| **Variance Components** | | | | |
| Individual | 0.94 (0.97) | 0.94 (0.97) | 0.94 (0.97) | 0.94 (0.97) |
| National**:** Country | 0.056 (0.24) | 0.043 (0.21) | 0.030 (0.17) | 0.029 (0.17) |
| Sex Female | | 0.014 (0.12) | 0.015 (0.12) | 0.01 (0.1) |
| **VPC** | 5.6% | 4.3% | 3.02% | 3% |

Model A baseline model to illustrate the between country variance. Model B includes sex female. Model C includes all country-level variables. Model D includes all cross-level interactions with sex female. Since the GGGI scale runs from 0-1 we multiply it by 10 so that the coefficient for GGGI represents a 0.1-point increase in the scale. GDP per capita is rescaled by dividing by 10,000, so that the coefficient represents the association with an increase of 1 x $10^4$ GDP per capita. Variance partition coefficient (VPC) indicates the proportion of unexplained variance attributable to differences between countries. *p<0.05 **p<0.01 ***p<0.001

### Figure S1: Forest plots of meta-analyses for each mental health outcome by regional sub-group

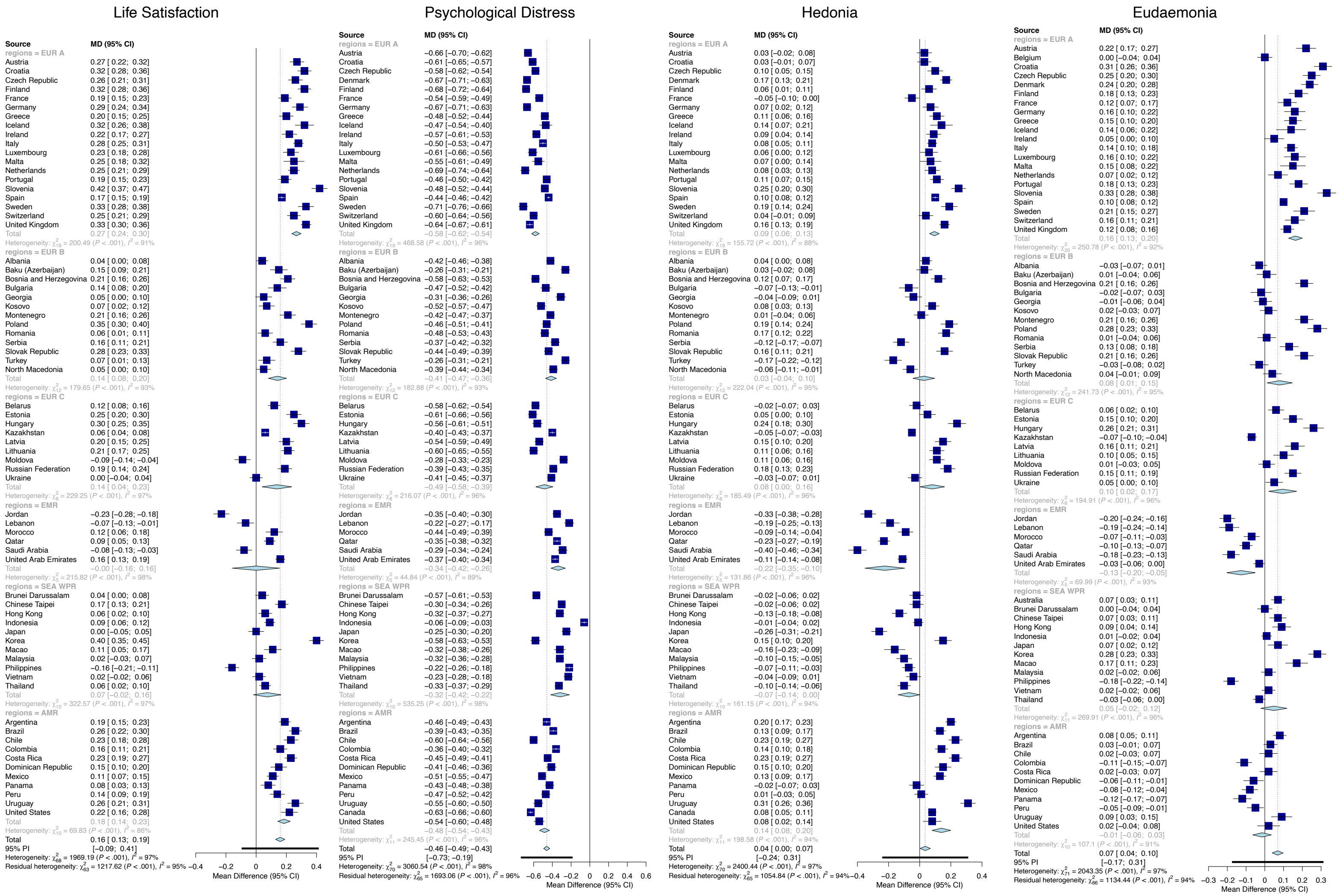

$I^2$ is the percentage of variation across nations due to heterogeneity rather than chance. MD = mean difference. High $I^2$ >85% indicates that there is considerable variation both within and between regions.

**Figure S2: Distributions of life satisfaction by gender and country**

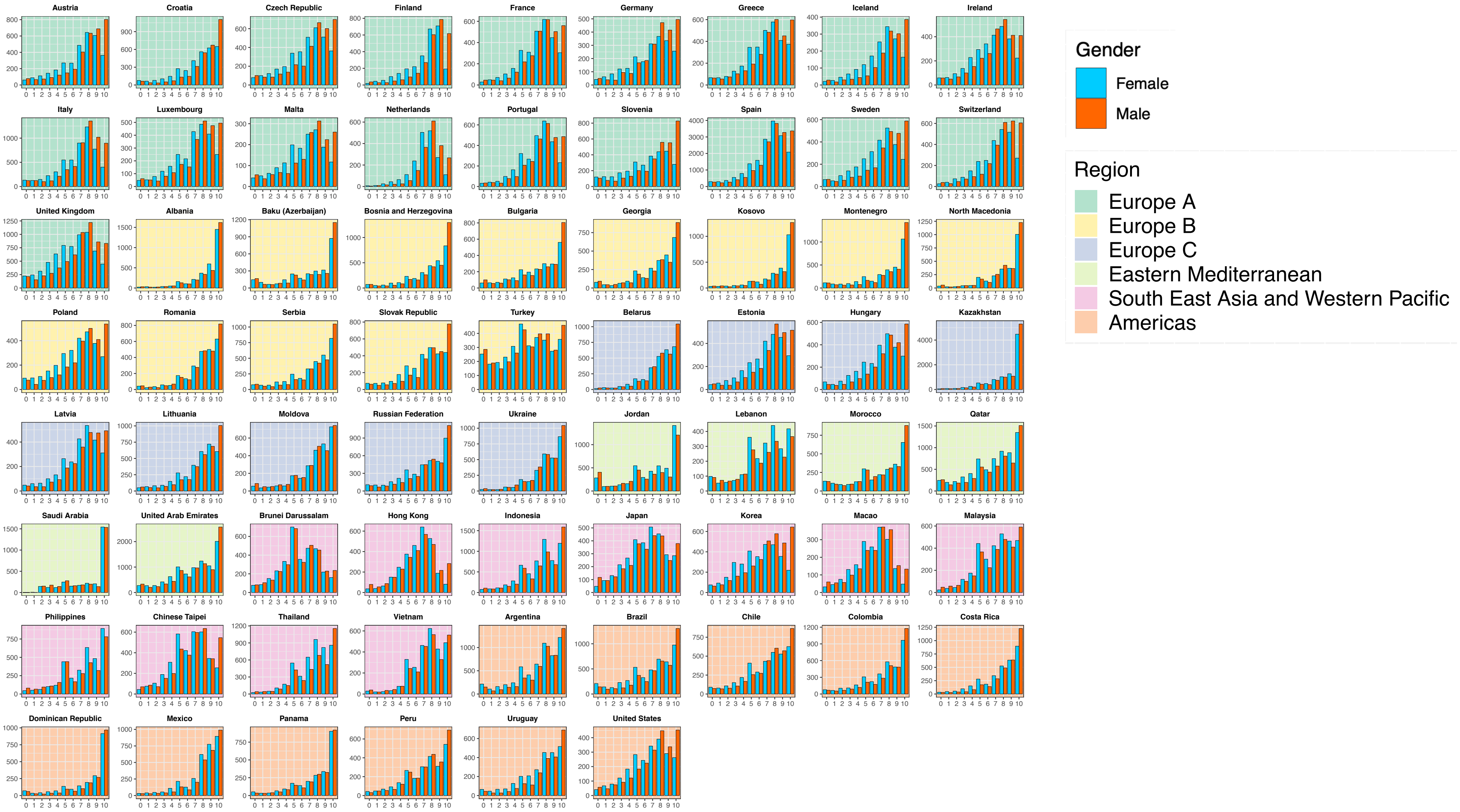

### Figure S3: Distributions of hedonia by gender and country

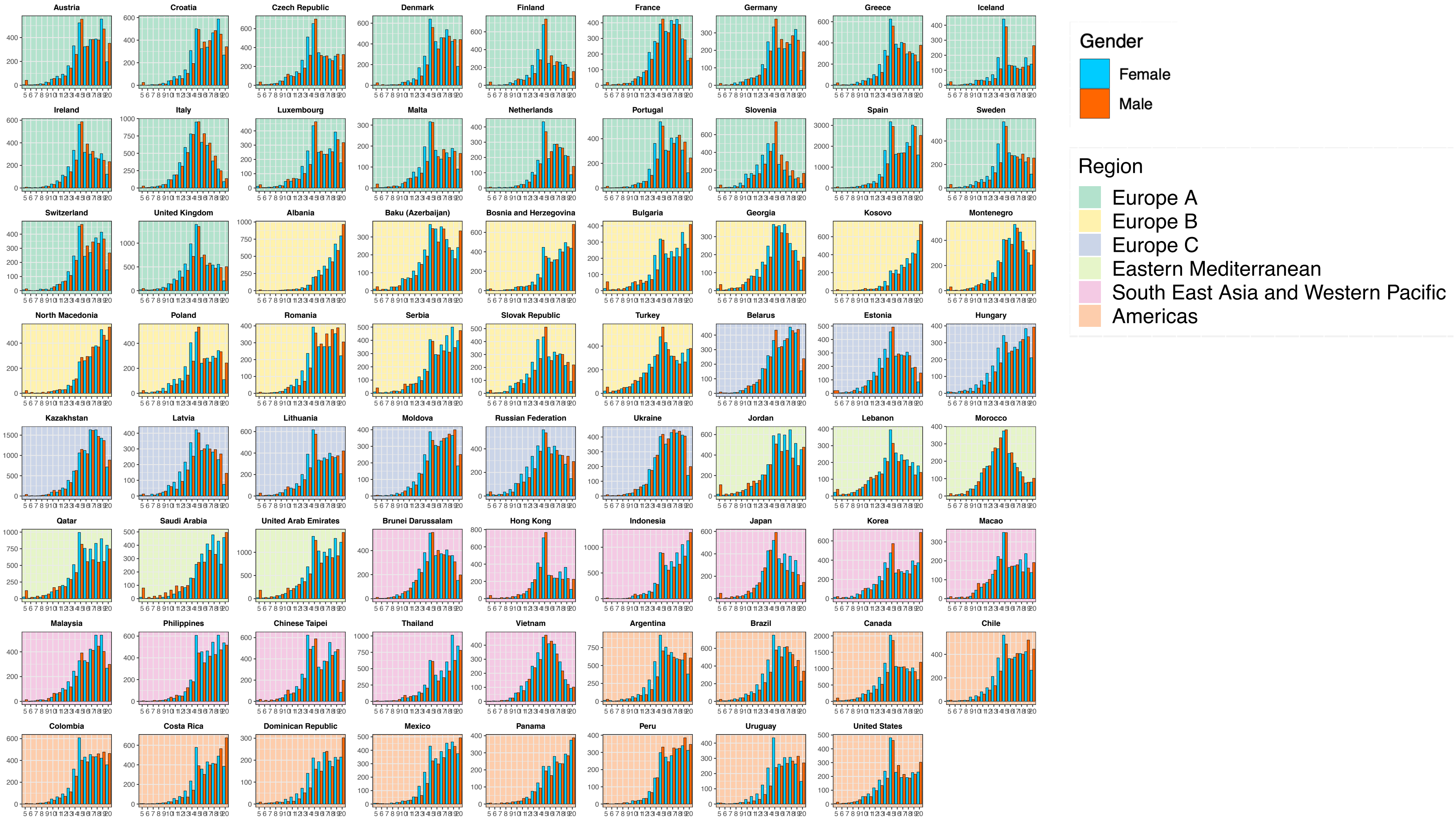

**Figure S4: Distributions of Eudaemonia by gender and country**

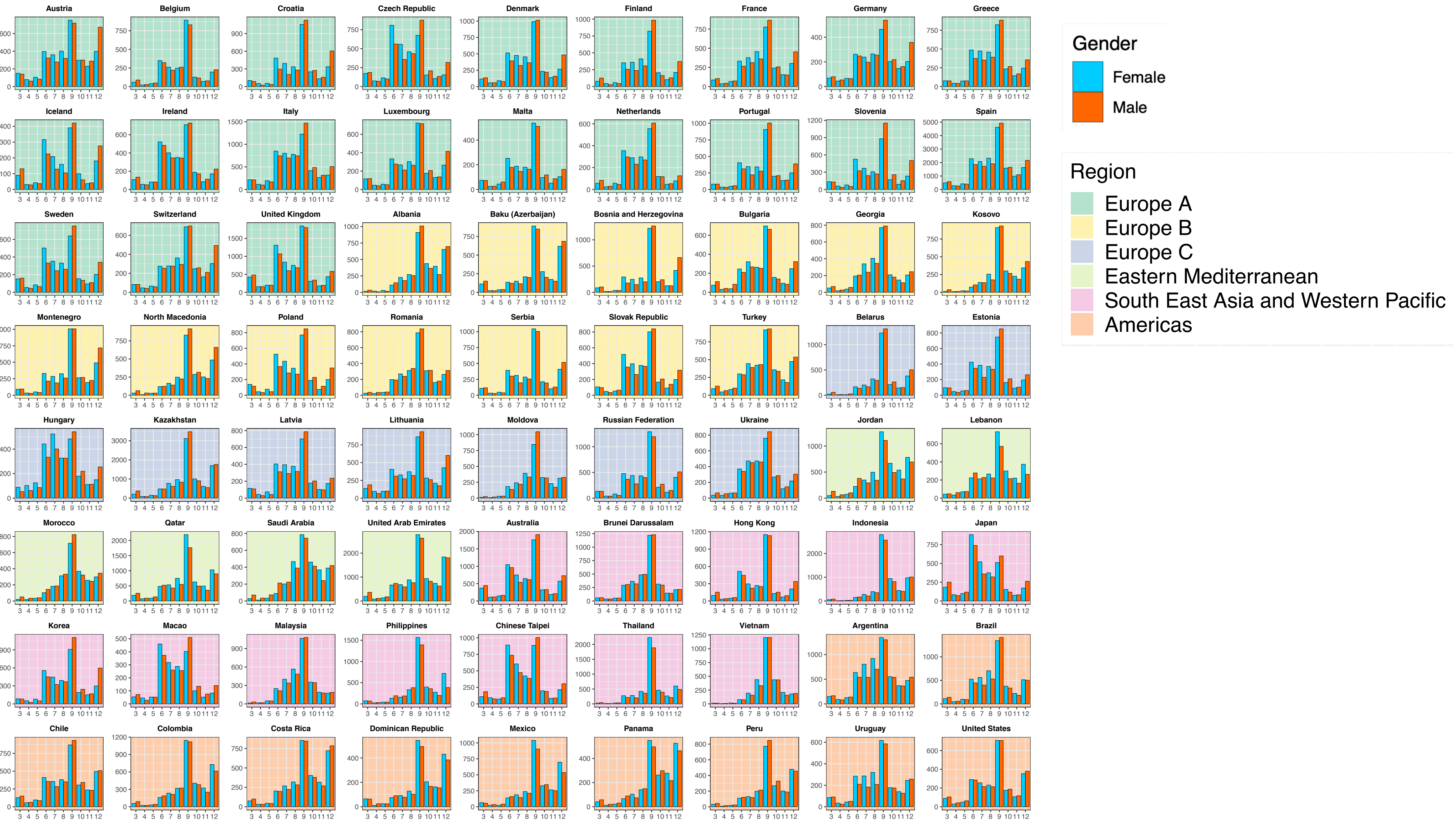

**Figure S5: Associations of country-level economic and gender equality indicators with average hedonia and eudaemonia by gender**

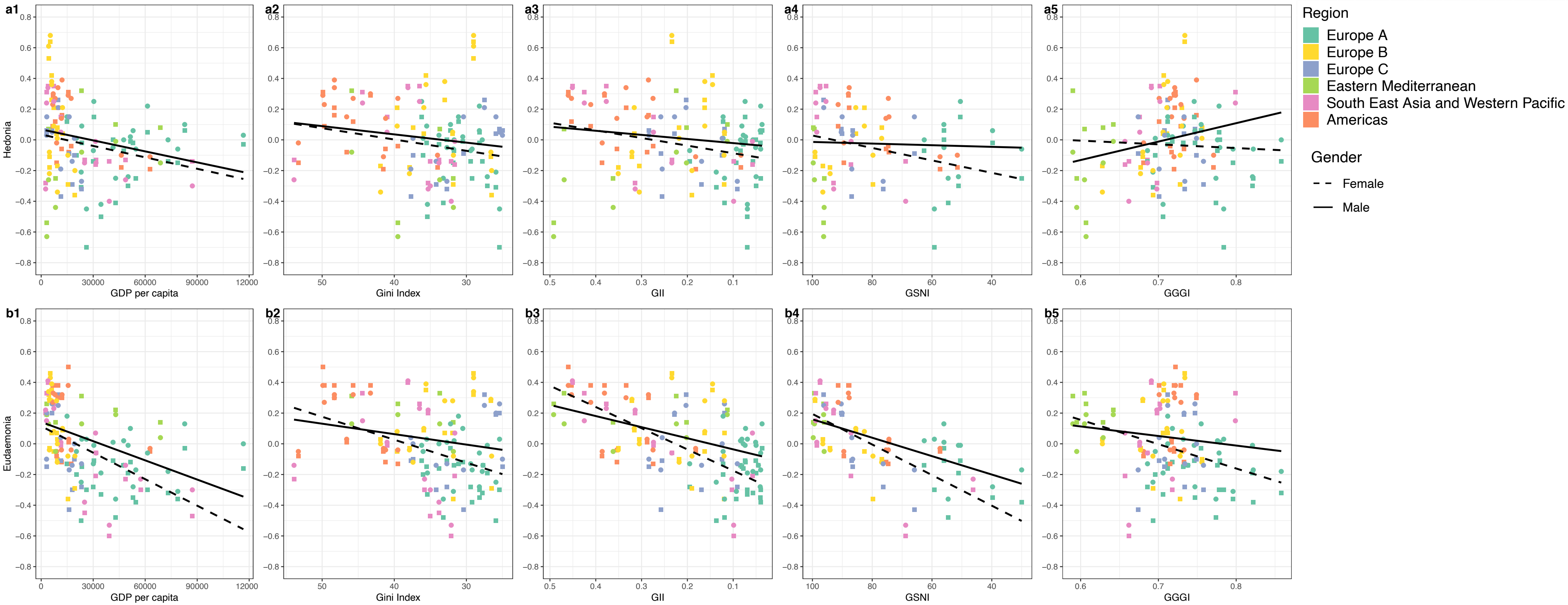

Country-level associations of economic indicators (GDP per capita and Gini) and gender equality indicators (GII, GSNI, and GGGI) with average standardised hedonia (a1-5) eudaemonia (b1-5) for females and males and coloured by region. The GII, GSNI and Gini scales are reversed so that they run from less equal to more equal permitting consistent interpretation with GGGI. A negative relationship indicates worse mental health across all outcomes. A larger distance between the regression lines indicates a larger gender gap. Abbreviations: Gini = income inequality, GII = gender inequality index, GSNI = gender social norms index, GGGI = global gender gap index.

**Figure S6: Associations of country-level economic and gender equality indicators with average life satisfaction and psychological distress by gender and with regional regression lines.**

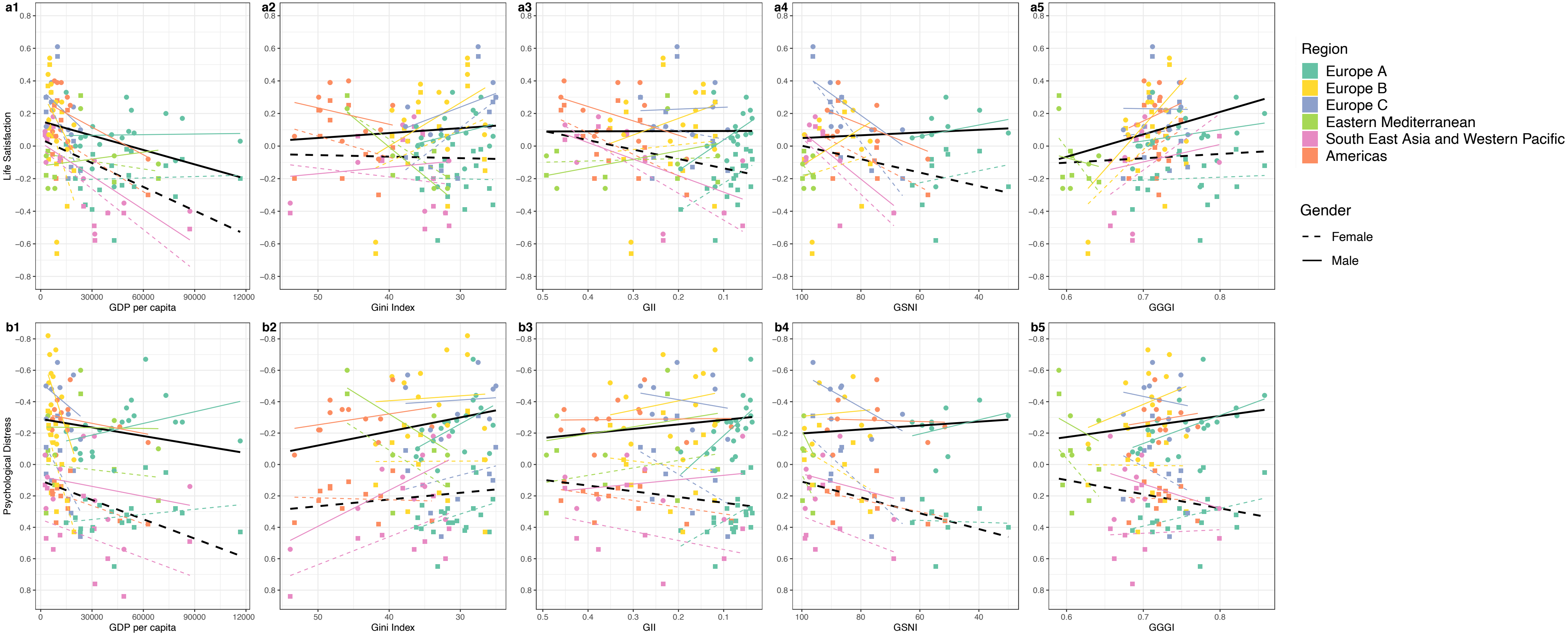

Country-level associations of economic indicators (GDP per capita and Gini) and gender equality indicators (GII, GSNI, and GGGI) with average standardised life satisfaction (a1-5) psychological distress (b1-5) for females and males and coloured by region. The GII, GSNI and Gini scales are reversed so that they run from less equal to more equal permitting consistent interpretation with GGGI. A negative relationship indicates worse mental health across all outcomes. A larger distance between the regression lines indicates a larger gender gap. Abbreviations: Gini = income inequality, GII = gender inequality index, GSNI = gender social norms index, GGGI = global gender gap index.
